# Predictors of Activity and Concentration Interference in Adults with Migraine: An Ecological Momentary Assessment Study

**DOI:** 10.64898/2026.10.11.26365249

**Authors:** Ali Bina, Golnaz Rezaee

## Abstract

**Objective:** To identify demographic and daily cognitive, affective, and contextual predictors of headache-related interference with activity and concentration in a smartphone-based migraine cohort, using statistical models appropriate to the ordinal structure of patient-reported interference.

**Background:** Migraine burden extends beyond headache to encompass cognitive and affective disturbance, yet most disability instruments summarize functioning retrospectively and cannot capture day-to-day variability in interference. Ecological momentary assessment addresses this limitation, yet few studies have examined cognitive and affective correlates of interference independent of pain intensity or have modeled interference using methods that respect its ordinal, non-equal-interval structure.

**Methods:** From May 2025 to April 2026, adult participants were enrolled in this prospective, decentralized observational study via specialty clinics and social media campaigns. Eligibility required fulfilling ICHD-3 diagnostic criteria for migraine using a validated electronic survey and experiencing between 3 and 20 monthly headache days. Enrolled individuals recorded daily smartphone diary entries and performed three brief cognitive tests over a 30-day monitoring period. Daily interference with activities and concentration were captured on 5-point ordinal scales and analyzed via cumulative link mixed-effects models incorporating participant-specific random intercepts. Predictor selection across nested specifications relied on likelihood ratio testing and Akaike Information Criterion (AIC) metrics, prioritizing the more parsimonious model when goodness-of-fit was comparable.

**Results:** The analytic sample comprised 1,267 headache-day observations from 143 participants. In the final models, brain fog (OR = 2.49 for activity, 3.19 for concentration), forgetfulness (OR = 1.48, 1.65), and mood disturbance (OR = 1.19, 1.31) were each independently associated with both outcomes (all p < 0.01), while age, sex, and employment status showed no significant association with either. Sleep quality (OR = 0.86) and anticipated next-day enjoyment (OR = 0.81) provided additional explanatory value for activity interference (p < 0.05) but not for concentration interference, for which the three-predictor model of brain fog, forgetfulness, and mood alone was retained as the most parsimonious final model.

**Conclusions:** Cognitive and affective symptoms, rather than demographic or socioeconomic characteristics, are the primary correlates of headache-related functional interference in this cohort. Activity and concentration interference are explained by partially distinct sets of daily influences, supporting the clinical value of assessing cognitive and affective symptoms alongside conventional pain-focused evaluation.

## Introduction

Migraine is a leading cause of disability worldwide, and its burden extends well beyond headache to encompass cognitive and affective disturbance, including impaired concentration, forgetfulness, and mood changes. These symptoms are increasingly recognized as clinically meaningful, treatment-relevant components of migraine, with cognitive complaints reported by a substantial proportion of individuals with migraine across large observational cohorts. Despite this recognition, cognitive and affective symptoms are rarely quantified as measurable outcomes in routine clinical care or treatment trials, which instead rely predominantly on pain-centered endpoints and on disability instruments, such as the Migraine Disability Assessment Scale, that summarize functioning retrospectively over the preceding weeks or months.

This retrospective approach carries a well-documented limitation: it cannot capture how functional interference varies from one headache day to the next, and it remains vulnerable to recall bias inherent to summarizing weeks of lived experience from memory. Ecological momentary assessment addresses this limitation directly, repeatedly sampling symptoms and context close in time to the experience itself, within a participant’s normal environment, rather than reconstructing them retrospectively. Prior ecological momentary assessment studies in migraine have characterized prodromal and interictal symptom clusters, including cognitive-affective functioning^1^, and have identified pain intensity, age, and allodynia as predictors of pain interference with daily activities^2^. Considerably less attention, however, has been directed toward whether cognitive and affective symptoms, independent of pain intensity itself, are associated with functional interference, and fewer studies still have modeled patient-reported interference using statistical approaches that respect its ordinal, non-equal-interval structure rather than treating it as a continuous measurement.

Parallel advances in computational biology and bioinformatics have increasingly demonstrated the value of high-dimensional, data-driven modeling in characterizing complex neuropsychiatric and neurological disease phenotypes^3-6^. In translational research, analytical frameworks ranging from multivariate feature extraction and machine-learning classification to molecular and clinical data integration have been vital for parsing phenotypic heterogeneity and identifying non-linear patterns within dense biomedical datasets^7-10^. Applying analogous computational rigor to high-frequency longitudinal digital health data—where real-time behavioral, clinical, and cognitive parameters are sampled continuously—bridges molecular insights with granular clinical manifestations. Leveraging robust statistical and computational modeling techniques to evaluate multi-feature ecological momentary data offers an objective path toward dissecting how cognitive and affective symptom dimensions uniquely contribute to day-level disability.

This study is a 30-day, smartphone-based, decentralized observational cohort study that paired daily symptom and contextual ratings with brief cognitive testing in adults with migraine and established the feasibility of this measurement approach in its baseline report^11^. A companion analysis of this cohort modeled brain fog and forgetfulness as outcomes rather than predictors and found substantially elevated odds of both symptoms on headache days relative to headache-free days, providing direct evidence that cognitive burden fluctuates meaningfully within this population at the day level^12^. This finding motivates a natural and, to date, unaddressed question: is this fluctuating cognitive and affective burden associated with greater functional interference on headache days, independent of pain intensity itself? The present study addresses this question using the study’s baseline and daily diary data.

Specifically, this study examined which demographic, clinical, and daily contextual characteristics were most strongly and consistently associated with headache-related interference with daily activity and concentration, modeled using cumulative link mixed models to account for the ordinal structure of both outcomes. We hypothesized, a priori, that cognitive symptoms, specifically brain fog and forgetfulness, together with mood disturbance, rather than demographic or socioeconomic characteristics, would constitute the dominant correlates of both outcomes. We further hypothesized that the relative contribution of contextual daily factors, including sleep quality and anticipated enjoyment, might differ between the two outcomes, given their differing conceptual proximity to cognitive functioning.

## Methods

### Study Design and Data Source

Participants were recruited from May 2025 to April 2026 through LinkedIn. Migraine diagnosis was assessed using a web-based, self-administered module based on ICHD-3 criteria and the validated American Migraine Prevalence and Prevention (AMPP) methodology. Diagnoses were not clinically confirmed. Eligibility criteria included age 18–60 years, smartphone ownership, English proficiency, migraine meeting screening criteria, and 3–20 headache days per month. Exclusion criteria included neurologic disorders other than migraine, chronic pain conditions, traumatic brain injury, severe psychiatric illness interfering with participation, and current alcohol or drug abuse.

We employed ecological momentary assessment (EMA), in which participants repeatedly reported on their symptoms and daily context in real time within their normal environment rather than relying on retrospective recall. Participants completed a baseline questionnaire at study enrollment, capturing demographic, clinical, and migraine-history information, and subsequently completed a brief once-daily EMA survey via a smartphone app over a 30-day monitoring period, assessing headache status, mood, sleep, cognitive symptoms, and functional interference on each day.

### Participants and Analytic Sample

Of the 169 participants who completed baseline assessments, 148 also contributed at least one daily diary entry and were included in this analysis. Because the two outcome variables of interest — pain interference with activity and pain interference with concentration — are recorded only on days when a participant answered “Yes” to the daily item “Since the last assessment, did you have headache?”, the analytic dataset was restricted to diary-days meeting this criterion, yielding 1,328 person-day observations from 147 participants. Baseline demographic covariates were then merged into the daily diary data by participant identifier. Observations missing a value on either outcome, any baseline covariate (age, sex, employment status), or any candidate daily predictor variable (brain fog, forgetfulness, mood, sleep hours, sleep quality, or anticipated stress or enjoyment of the following day) evaluated in the regression models were excluded via listwise deletion (61 observations from 4 additional participants), resulting in a final analytic sample of 1,267 person-day observations from 143 participants.

### Migraine Frequency Classification

Participants were classified at baseline into three migraine-frequency groups based on self-reported monthly headache days (MHD): Low-Frequency Episodic Migraine (LFEM, 2–7 headache days per month), High-Frequency Episodic Migraine (HFEM, 8–14 headache days per month), and Chronic Migraine (CM, ≥15 headache days per month)^11^. Migraine-frequency group was used to describe and compare baseline characteristics of the analytic sample (Table 1); it was not entered as a predictor in the regression models, since MHD is a retrospective, monthly baseline measure, whereas the regression models examine same-day, prospectively assessed predictors of daily functional interference.

**Table 1.** Baseline Characteristics by Migraine-Frequency Group.

| Characteristic | Overall | Low Frequency EM | High Frequency EM | Chronic Migraine | P |
| --- | --- | --- | --- | --- | --- |
| N | 143 (100%) | 44 (30.8%) | 30 (21.0%) | 69 (48.3%) |  |
| Age | 37.86 (11.38) | 38.91 (12.67) | 36.20 (9.43) | 37.91 (11.36) | 0.606 |
| Sex: Female | 116 (81.1%) | 32 (72.7%) | 25 (83.3%) | 59 (85.5%) | 0.225 |
| Sex: Male | 27 (18.9%) | 12 (27.3%) | 5 (16.7%) | 10 (14.5%) | 0.225 |
| Race: Asian | 4 (2.8%) | 3 (6.8%) | 0 (0.0%) | 1 (1.4%) | 0.236 |
| Race: Black or African American | 18 (12.6%) | 5 (11.4%) | 7 (23.3%) | 6 (8.7%) | 0.236 |
| Race: Two or More Races | 5 (3.5%) | 2 (4.5%) | 1 (3.3%) | 2 (2.9%) | 0.236 |
| Race: White | 116 (81.1%) | 34 (77.3%) | 22 (73.3%) | 60 (87.0%) | 0.236 |
| Hispanic: No | 136 (95.1%) | 42 (95.5%) | 29 (96.7%) | 65 (94.2%) | 1.000 |
| Hispanic: Yes | 7 (4.9%) | 2 (4.5%) | 1 (3.3%) | 4 (5.8%) | 1.000 |
| Marital Status: Divorced | 4 (2.8%) | 1 (2.3%) | 0 (0.0%) | 3 (4.3%) | 0.917 |
| Marital Status: Living with Partner | 19 (13.3%) | 8 (18.2%) | 4 (13.3%) | 7 (10.1%) | 0.917 |
| Marital Status: Married | 72 (50.3%) | 19 (43.2%) | 16 (53.3%) | 37 (53.6%) | 0.917 |
| Marital Status: Prefer Not to Answer | 1 (0.7%) | 0 (0.0%) | 0 (0.0%) | 1 (1.4%) | 0.917 |
| Marital Status: Separated | 2 (1.4%) | 1 (2.3%) | 0 (0.0%) | 1 (1.4%) | 0.917 |
| Marital Status: Single | 45 (31.5%) | 15 (34.1%) | 10 (33.3%) | 20 (29.0%) | 0.917 |
| Income: Prefer Not to Answer | 29 (20.3%) | 8 (18.2%) | 5 (16.7%) | 16 (23.2%) | 0.030 |
| Income: \$25,000-50,000 | 18 (12.6%) | 12 (27.3%) | 3 (10.0%) | 3 (4.3%) | 0.030 |
| Income: \$50,000-75,000 | 13 (9.1%) | 6 (13.6%) | 1 (3.3%) | 6 (8.7%) | 0.030 |
| Income: \$75,000-100,000 | 21 (14.7%) | 5 (11.4%) | 4 (13.3%) | 12 (17.4%) | 0.030 |
| Income: >\$100,000 | 62 (43.4%) | 13 (29.5%) | 17 (56.7%) | 32 (46.4%) | 0.030 |
| Education: College Degree or More | 120 (83.9%) | 42 (95.5%) | 24 (80.0%) | 54 (78.3%) | 0.083 |
| Education: High School to Some College | 20 (14.0%) | 2 (4.5%) | 5 (16.7%) | 13 (18.8%) | 0.083 |
| Education: High School or Less | 3 (2.1%) | 0 (0.0%) | 1 (3.3%) | 2 (2.9%) | 0.083 |
| Employment: Full Time | 83 (58.0%) | 28 (63.6%) | 19 (63.3%) | 36 (52.2%) | 0.334 |
| Employment: Part Time | 23 (16.1%) | 9 (20.5%) | 2 (6.7%) | 12 (17.4%) | 0.334 |
| Employment: Homemaker | 7 (4.9%) | 2 (4.5%) | 2 (6.7%) | 3 (4.3%) | 0.334 |
| Employment: Disability | 9 (6.3%) | 1 (2.3%) | 1 (3.3%) | 7 (10.1%) | 0.334 |
| Employment: Looking for Work | 8 (5.6%) | 2 (4.5%) | 4 (13.3%) | 2 (2.9%) | 0.334 |
| Employment: Not Employed | 3 (2.1%) | 1 (2.3%) | 1 (3.3%) | 1 (1.4%) | 0.334 |
| Employment: Retired | 4 (2.8%) | 0 (0.0%) | 0 (0.0%) | 4 (5.8%) | 0.334 |
| Employment: Student | 6 (4.2%) | 1 (2.3%) | 1 (3.3%) | 4 (5.8%) | 0.334 |
| Sleep | 7.03 (1.15) | 6.93 (1.15) | 7.20 (1.16) | 7.03 (1.16) | 0.619 |
| Pain | 8.29 (1.53) | 8.09 (1.61) | 8.00 (1.82) | 8.54 (1.31) | 0.166 |
| MSSS | 18.21 (2.38) | 18.14 (2.38) | 17.57 (2.69) | 18.54 (2.21) | 0.172 |
| Allodynia: Never | 15 (10.5%) | 7 (15.9%) | 1 (3.3%) | 7 (10.1%) | 0.040 |
| Allodynia: Rarely | 32 (22.4%) | 13 (29.5%) | 8 (26.7%) | 11 (15.9%) | 0.040 |
| Allodynia: <Half the Time | 45 (31.5%) | 13 (29.5%) | 14 (46.7%) | 18 (26.1%) | 0.040 |
| Allodynia: ≥Half the Time | 51 (35.7%) | 11 (25.0%) | 7 (23.3%) | 33 (47.8%) | 0.040 |
| Aura: Never | 30 (21.0%) | 10 (22.7%) | 6 (20.0%) | 14 (20.3%) | 0.644 |
| Aura: Rarely | 32 (22.4%) | 9 (20.5%) | 10 (33.3%) | 13 (18.8%) | 0.644 |
| Aura: <Half the Time | 42 (29.4%) | 11 (25.0%) | 9 (30.0%) | 22 (31.9%) | 0.644 |
| Aura: ≥Half the Time | 39 (27.3%) | 14 (31.8%) | 5 (16.7%) | 20 (29.0%) | 0.644 |
| MIDAS | 53.50 [32.00-103.75] | 36.00 [20.75-51.25] | 51.50 [33.50-75.75] | 93.50 [50.00-139.75] | <0.001 |
| MIDAS Category: None | 5 (3.6%) | 2 (4.7%) | 1 (3.3%) | 2 (3.1%) | 0.027 |
| MIDAS Category: Mild | 5 (3.6%) | 2 (4.7%) | 0 (0.0%) | 3 (4.6%) | 0.027 |
| MIDAS Category: Moderate | 7 (5.1%) | 6 (14.0%) | 1 (3.3%) | 0 (0.0%) | 0.027 |
| MIDAS Category: Severe | 121 (87.7%) | 33 (76.7%) | 28 (93.3%) | 60 (92.3%) | 0.027 |
| Concentration Interference Days | 10.00 [5.50-20.00] | 5.00 [4.00-8.00] | 11.00 [8.00-15.00] | 15.00 [10.00-20.00] | <0.001 |
| Functional Interference Days | 10.00 [6.00-20.00] | 6.00 [4.00-10.00] | 10.00 [8.00-10.00] | 20.00 [15.00-25.00] | <0.001 |
| MIBS | 5.00 [4.00-6.00] | 4.00 [4.00-5.75] | 4.00 [4.00-6.00] | 5.00 [4.00-7.00] | 0.202 |
| MIBS Category: None | 1 (0.7%) | 0 (0.0%) | 1 (3.3%) | 0 (0.0%) | 0.117 |
| MIBS Category: Mild | 8 (5.8%) | 2 (4.7%) | 1 (3.3%) | 5 (7.7%) | 0.117 |
| MIBS Category: Moderate | 114 (82.6%) | 40 (93.0%) | 24 (80.0%) | 50 (76.9%) | 0.117 |
| MIBS Category: Severe | 15 (10.9%) | 1 (2.3%) | 4 (13.3%) | 10 (15.4%) | 0.117 |
| PHQ4 | 4.14 (2.84) | 3.39 (2.54) | 4.13 (2.94) | 4.62 (2.92) | 0.078 |
| PHQ4 Category: None | 41 (28.7%) | 15 (34.1%) | 10 (33.3%) | 16 (23.2%) | 0.630 |
| PHQ4 Category: Mild | 62 (43.4%) | 21 (47.7%) | 11 (36.7%) | 30 (43.5%) | 0.630 |
| PHQ4 Category: Moderate | 27 (18.9%) | 6 (13.6%) | 6 (20.0%) | 15 (21.7%) | 0.630 |
| PHQ4 Category: Severe | 13 (9.1%) | 2 (4.5%) | 3 (10.0%) | 8 (11.6%) | 0.630 |
| PSS4 | 13.00 [12.00-14.00] | 12.00 [12.00-13.00] | 12.50 [12.00-14.00] | 13.00 [12.00-14.00] | 0.804 |

**Note:** LFEM = Low-Frequency Episodic Migraine; HFEM = High-Frequency Episodic Migraine; CM = Chronic Migraine; MSSS = Migraine Symptom Severity Score; MIDAS = Migraine Disability Assessment Score; MIBS = Migraine Interictal Burden Scale; PHQ-4 = Patient Health Questionnaire-4; PSS-4 = Perceived Stress Scale-4. Continuous variables are presented as mean (SD) or median [Q1–Q3] depending on distribution (see Methods); categorical variables are presented as n (%). PSS-4 has no established categorical severity classification and is therefore reported as a continuous score only. Reported p-values reflect omnibus comparisons across all three migraine-frequency groups (ANOVA, Kruskal-Wallis, chi-square, or Fisher's exact test with Monte Carlo simulation, as appropriate); pairwise comparisons for significant characteristics are reported in the Results text. The Hispanic ethnicity subgroup was small (n = 7); its distribution across migraine-frequency groups closely matched the counts expected under no association, yielding a simulated Fisher's exact p-value at the upper bound (1.000).

### Outcome Variables

Two outcome variables were assessed on each headache-day, defined as a diary-day on which the participant reported a headache since the last assessment. This affirmative response triggered a block of headache-episode follow-up questions within the same daily survey, including the two items of interest for this analysis.

The first, “How much did pain interfere with your daily activities?”, captured pain interference with activity, while the second, “How much pain interfered with your ability to concentrate?”, captured pain interference with concentration. Both items used an identical five-level ordinal response format — Not at all, A little bit, Moderately, Quite a bit, and Extremely — completed within the same daily survey submission. Both outcomes were analyzed in their native five-category ordinal form using cumulative link mixed models (CLMMs), given the unequal, data-dependent spacing between adjacent response categories; this modeling approach is described in full in the Statistical Analysis subsection.

### Predictor Variables

Three baseline demographic and socioeconomic characteristics, collected at study enrollment, were included as candidate predictors: age (years, continuous), sex (Female/Male), and employment status. Employment status was originally recorded across nine categories: employed full-time, employed part-time, student, retired, homemaker, long-term or short-term disability, looking for a job, not employed and not looking for a job, and prefer not to answer. Because several of these categories contained very few participants, retaining the full nine-level variable in the regression models risked unstable coefficient estimates for the smallest groups. Employment status was therefore collapsed into a binary indicator — Employed (full- or part-time) versus Not Employed (student, retired, homemaker, disability, or unemployed, whether or not currently seeking work) — which preserves a clinically interpretable distinction while avoiding sparse-category estimation problems. Participants who selected “Prefer not to answer” for employment status were treated as missing on this variable, since it does not correspond to a genuine employment status.

Seven daily-level predictors, assessed once per diary-day regardless of whether a headache was reported since the last assessment, were evaluated as candidate predictors of same-day pain interference. Brain fog severity was assessed with the item “Did you feel that you have a ‘brain fog’ and are not as sharp as usual today?”, and mood disturbance with “Have you been feeling moody today?”; both used an identical five-level ordinal response format (Not at all, Slightly, Somewhat, Very much, Extremely), scored 0–4. Forgetfulness was assessed with the binary item “Did you forget any tasks today?” (Yes/No). The remaining four daily contextual predictors comprised sleep, assessed with two separate items — sleep duration, a continuous measure of hours slept the previous night, and sleep quality, assessed with “How was the quality of your sleep last night?” using a five-level ordinal format specific to sleep (Very Poor, Poor, Fair, Good, Very Good), scored 0–4 — and two forward-looking items asking participants to rate their anticipated stress and enjoyment of the following day, each using its own five-level ordinal response format and scored 0–4. All seven daily predictors were treated as numeric (continuous or 0–4 ordinal-scored) or binary variables in the regression models; the ordinal treatment applied in this study’s cumulative link mixed models concerns only the outcome variables, not the predictors, even where a predictor is itself a Likert-type item.

### Statistical Analysis

All statistical analyses were conducted in R version 4.6.1 (R Core Team, 2026), with a fixed random seed (123) set prior to any simulation-based procedure to ensure reproducibility. Baseline characteristics of the analytic sample were summarized overall and stratified by baseline migraine-frequency group (LFEM, HFEM, CM) in Table 1. Continuous variables were tested for normality using the Shapiro-Wilk test; normally distributed variables are reported as mean (SD) and compared across groups using one-way analysis of variance (ANOVA), while non-normally distributed variables are reported as median [Q1–Q3] and compared using the Kruskal-Wallis test^13, 14^. Categorical variables are reported as n (%) and compared using the chi-square test, or Fisher’s exact test with Monte Carlo simulation of the p-value when any expected cell count was below 5^15^. Because each comparison spans three migraine-frequency groups, reported p-values test the omnibus null hypothesis that at least one group differs from the others, rather than identifying which specific pair of groups differs. For characteristics with a significant omnibus test (p < 0.05), post-hoc pairwise comparisons were performed using Dunn’s test with Bonferroni correction (dunn.test package, version 1.4.1)^16^. A two-sided α of 0.05 was used to define statistical significance throughout.

Both outcome variables were modeled using cumulative link mixed models (CLMMs; mixed-effects proportional-odds logistic regression)^17^. This approach was used in preference to linear mixed-effects models because both outcomes are ordinal with unequal spacing between adjacent response categories, which a CLMM estimates directly from the data rather than assuming. All models specified a logit link function and included a random intercept for participant to account for the non-independence of repeated within-person observations. Base models containing only the three baseline predictors did not reliably converge under default settings and were refit with adjusted convergence control parameters (maximum iterations = 1000, gradient tolerance = 1×10 □ □).

For each outcome, a nested sequence of eight models (Models 1–8) evaluated the individual and joint contribution of three daily predictors identified a priori as the primary hypothesized correlates of functional interference — brain fog, forgetfulness, and mood — each added to a base model containing age, sex, and employment status. Model 1 comprised the base model alone; Models 2–4 introduced each symptom predictor individually; Models 5–7 introduced each possible pair; and Model 8 combined all three.

A second, exploratory sequence (Models 9–15) examined four daily contextual variables — sleep hours, sleep quality, anticipated stress, and anticipated enjoyment — added individually to the same base model (Models 9–12). Notably, all seven daily predictors examined in this study, the three primary predictors as well as these four contextual variables, reached statistical significance when tested individually. Because sleep hours and sleep quality, and anticipated stress and anticipated enjoyment, represent conceptually related pairs, each pair was subsequently modeled jointly (Models 13–14) before all four contextual variables were combined into a single model (Model 15). Within each pair, sleep quality and anticipated enjoyment showed the stronger, more consistent association with the outcomes, relative to sleep hours and anticipated stress, respectively. A further model (Model 16) retained only this reduced two-variable contextual set — sleep quality and anticipated enjoyment — alongside the base model, omitting sleep hours and anticipated stress, to test whether this reduction resulted in a significant loss of fit relative to Model 15.

The two predictor sets were then combined. Model 17 incorporated all seven daily predictors alongside the base model, while Model 18 combined the reduced contextual set (sleep quality and anticipated enjoyment) with the three primary predictors (brain fog, forgetfulness, and mood). All 18 fitted models for each outcome, including regression coefficients, significance levels, and AIC, are reported in full in Table 2.

**Table 2.**
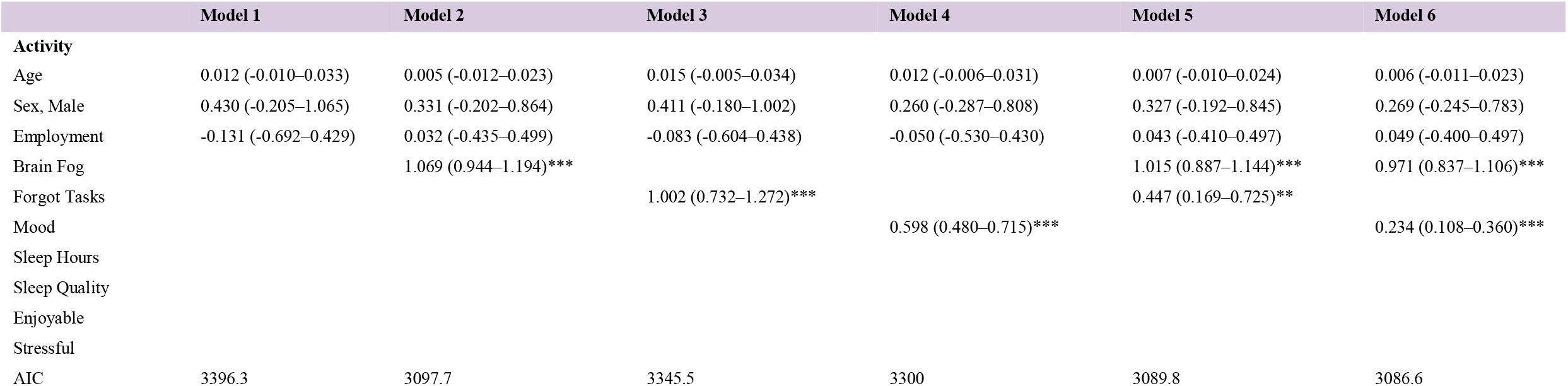
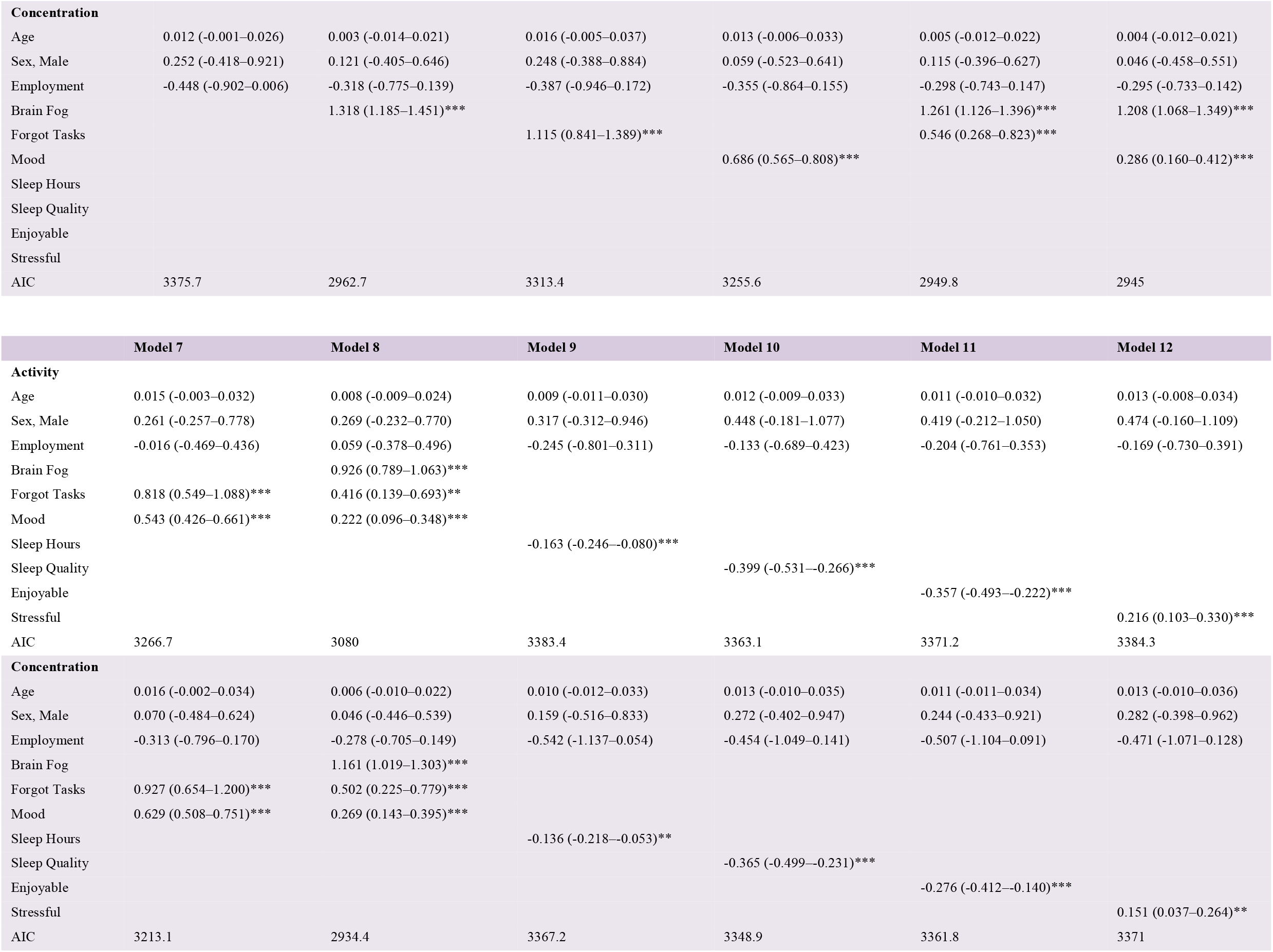
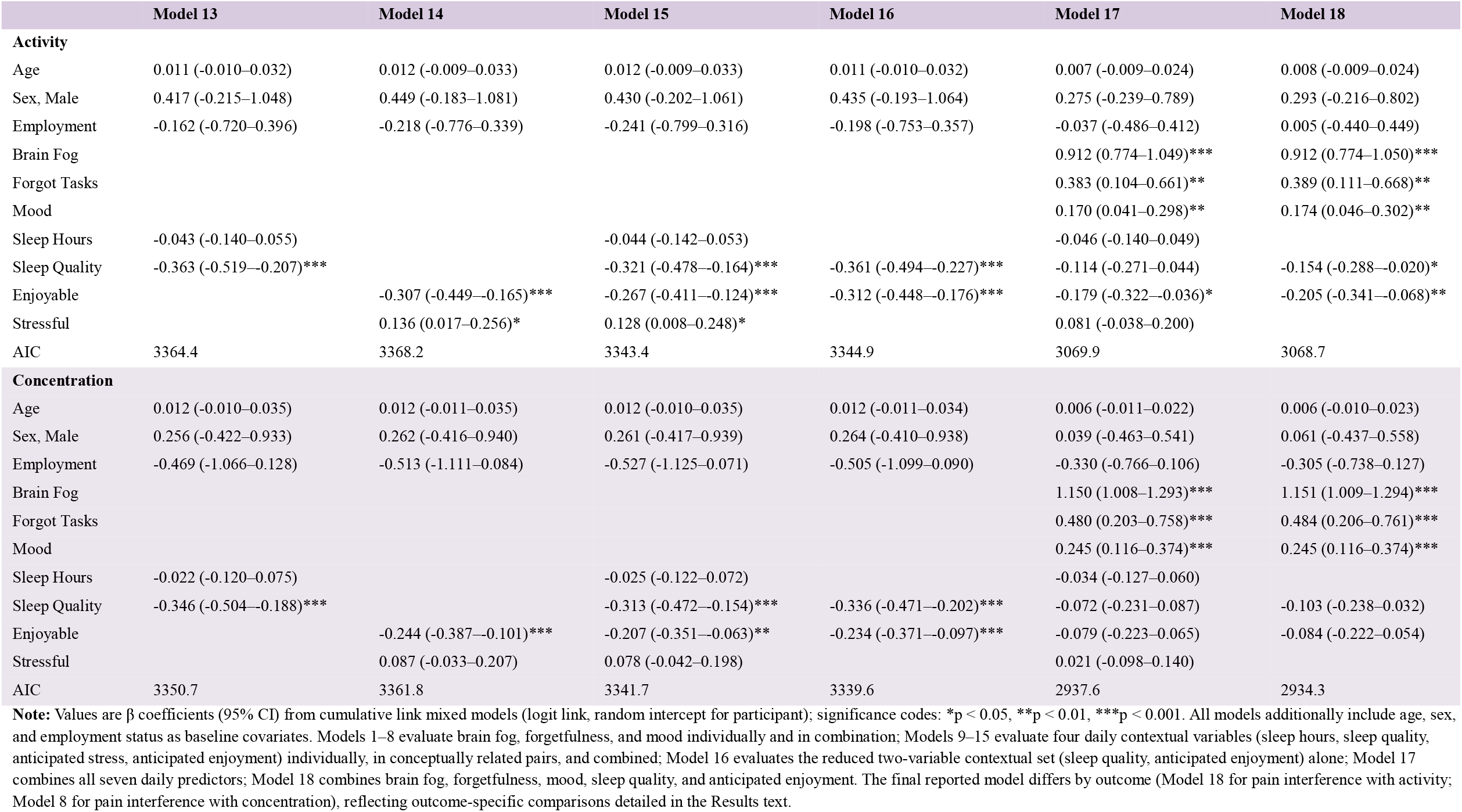
Cumulative Link Mixed Model Results (Models 1–6)

Model fit was compared using the Akaike Information Criterion (AIC), with lower values indicating better fit relative to model complexity. Because a lower AIC alone does not establish that a more complex model significantly improves upon a simpler one, comparisons between models were further evaluated using likelihood ratio tests (LRTs), which formally test whether the observed improvement in fit exceeds what would be expected by chance. For each outcome, model comparisons proceeded in two stages. First, within the contextual-variable sequence, Model 16 (the reduced two-variable contextual set) was compared against Model 15 (all four contextual variables) to confirm that omitting sleep hours and anticipated stress did not significantly reduce fit. Second, Model 17 (all seven daily predictors) and Model 18 (the reduced contextual set combined with brain fog, forgetfulness, and mood) were each compared against Model 8 (brain fog, forgetfulness, and mood only), and against each other, to determine whether contextual variables meaningfully improved prediction beyond the primary predictor set. Where an LRT indicated no significant difference between two models, the model with the lower AIC was preferred, reflecting comparable fit achieved with fewer parameters. The final reported model for each outcome was the model achieving the lowest AIC among all statistically justified candidates; because the contribution of contextual variables differed between outcomes (see Results), the final model differs between the two outcomes.

## Results

### Sample Characteristics

The analytic sample comprised 143 participants, classified at baseline into three migraine-frequency groups: 44 (30.8%) with Low-Frequency Episodic Migraine, 30 (21.0%) with High-Frequency Episodic Migraine, and 69 (48.3%) with Chronic Migraine. The overall sample had a mean age of 37.86 years (SD = 11.38) and was predominantly female (81.1%). Full baseline demographic and clinical characteristics, stratified by migraine-frequency group, are presented in Table 1.

Across the three migraine-frequency groups, most demographic characteristics were well balanced, including age, sex, race, ethnicity, marital status, education, and employment status (all p > 0.05). Household income was the only demographic characteristic to differ significantly across groups (p = 0.030), with a notably lower proportion of participants in the Chronic Migraine group reporting income between $25,000 and $50,000 (4.3%) relative to the Low-Frequency Episodic group (27.3%).

By contrast, several clinical and headache-related disability characteristics differed significantly by migraine-frequency group. Allodynia frequency differed significantly across groups (p = 0.040), with a substantially higher proportion of the Chronic Migraine group reporting allodynia at least half the time (47.8%) relative to the Low- and High-Frequency Episodic groups (25.0% and 23.3%, respectively). Headache-related disability, as measured by the MIDAS total score^18^, differed significantly across groups (p < 0.001), as did the corresponding four-level MIDAS disability category (p = 0.027); because these two variables represent a continuous score and its derived clinical classification, respectively, they were tested independently and are not expected to produce identical results. Functional interference also differed significantly across groups, both in days of headache-related interference with concentration, memory, or clear thinking (p < 0.001) and in days of headache-related interference with work, study, or enjoyment of life (p < 0.001). No significant between-group differences were observed for sleep hours, pain intensity, MSSS total score^19^, MIBS total score or category ^20^, PHQ-4 total score or category^21^, or PSS-4^22^ total score (all p > 0.05).

Post-hoc pairwise comparisons using Dunn’s test with Bonferroni correction indicated that MIDAS total score differed significantly between all three migraine-frequency groups (Low-vs. High-Frequency Episodic, p = 0.040; Low-Frequency Episodic vs. Chronic Migraine, p < 0.001; High-Frequency Episodic vs. Chronic Migraine, p = 0.024), consistent with a stepwise increase in disability with increasing headache frequency. Days of headache-related interference with work, study, or enjoyment of life differed significantly between the Chronic Migraine group and both episodic groups (Low-Frequency Episodic vs. Chronic Migraine, p < 0.001; High-Frequency Episodic vs. Chronic Migraine, p < 0.001) but not between the Low- and High-Frequency Episodic groups (p = 0.062). Days of headache-related interference with concentration, memory, or clear thinking showed a distinct pattern: the Low-Frequency Episodic group differed significantly from both the High-Frequency Episodic and Chronic Migraine groups (both p < 0.001), which did not differ significantly from each other (p = 0.219), suggesting that interference with concentration is already substantially elevated at high headache frequency, without further significant increase at chronic-level frequency.

### Predictors of Pain Interference with Activity

Across the eight-model sequence evaluating brain fog, forgetfulness, and mood (Models 1–8), model fit improved substantially with the addition of each predictor. The base model containing only age, sex, and employment status (Model 1, AIC = 3396.3) showed the poorest fit of the sequence. Brain fog was the strongest individual predictor (Model 2, AIC = 3097.7), followed by forgetfulness (Model 3, AIC = 3345.5) and mood (Model 4, AIC = 3300.0). The model including all three predictors jointly (Model 8) achieved the best fit within this sequence (AIC = 3080.0), with brain fog (β = 0.93, p < 0.001), forgetfulness (β = 0.42, p = 0.003), and mood (β = 0.22, p < 0.001) each independently and significantly associated with greater odds of activity interference.

All four daily contextual variables were individually significant predictors of activity interference when tested alone (Models 9–12, all p < 0.05). Sleep quality showed a better individual fit than sleep hours (Model 10, AIC = 3363.1 vs. Model 9, AIC = 3383.4), and anticipated enjoyment showed a better individual fit than anticipated stress (Model 11, AIC = 3371.2 vs. Model 12, AIC = 3384.3). This pattern was reinforced when each pair was modeled jointly: sleep hours’ association was fully attenuated once sleep quality was included (Model 13, AIC = 3364.4; sleep hours β = −0.04, p > 0.05; sleep quality β = −0.36, p < 0.001), and anticipated stress’s association weakened, though it remained significant, once anticipated enjoyment was included (Model 14, AIC = 3368.2; stress β = 0.14, p < 0.05; enjoyment β = −0.31, p < 0.001). A model combining all four contextual variables (Model 15, AIC = 3343.4) improved fit somewhat beyond either paired model. A reduced model retaining only sleep quality and anticipated enjoyment (Model 16, AIC = 3344.9) showed no significant loss of fit relative to Model 15 (*x*^2^ = 5.47, df = 2, p = 0.065), indicating that sleep hours and anticipated stress contributed little unique information beyond sleep quality and anticipated enjoyment, even without brain fog, forgetfulness, or mood in the model.

Combining both predictor sets, a likelihood ratio test comparing Model 17 (all seven daily predictors, AIC = 3069.9) against Model 8 indicated that the contextual variables significantly improved model fit (*x*^2^ = 18.10, df = 4, p = 0.001). A further likelihood ratio test comparing Model 18 (the reduced contextual set combined with brain fog, forgetfulness, and mood, AIC = 3068.7) against Model 8 confirmed a significant improvement (*x*^2^ = 15.26, df = 2, p < 0.001), while comparison of Model 17 against Model 18 showed no significant difference in fit (*x*^2^ = 2.84, df = 2, p = 0.242). Model 18 achieved the lowest AIC among all candidate models for this outcome and was retained as the final model.

In the final model (Model 18), brain fog (β = 0.91, SE = 0.07, OR = 2.49, p < 0.001), forgetfulness (β = 0.39, SE = 0.14, OR = 1.48, p = 0.006), mood (β = 0.17, SE = 0.07, OR = 1.19, p = 0.008), sleep quality (β = −0.15, SE = 0.07, OR = 0.86, p = 0.024), and anticipated enjoyment (β = −0.20, SE = 0.07, OR = 0.81, p = 0.003) were each independently and significantly associated with pain interference with activity, while age, sex, and employment status showed no significant association.

### Predictors of Pain Interference with Concentration

The same 18-model sequence was fit for pain interference with concentration. The base model (Model 1, AIC = 3375.7) again showed the poorest fit. Brain fog was the strongest individual predictor (Model 2, AIC = 2962.7), followed by forgetfulness (Model 3, AIC = 3313.4) and mood (Model 4, AIC = 3255.6). The model including all three predictors jointly (Model 8) achieved the best fit within this sequence (AIC = 2934.4), with brain fog (β = 1.16, p < 0.001), forgetfulness (β = 0.50, p < 0.001), and mood (β = 0.27, p < 0.001) each independently and significantly associated with greater odds of concentration interference.

All four daily contextual variables were individually significant predictors of concentration interference when tested alone (Models 9–12, all p < 0.05). As with the activity outcome, sleep quality showed a better individual fit than sleep hours (Model 10, AIC = 3348.9 vs. Model 9, AIC = 3367.2), and anticipated enjoyment showed a better individual fit than anticipated stress (Model 11, AIC = 3361.8 vs. Model 12, AIC = 3371.0). When each pair was modeled jointly, sleep hours’ association was fully attenuated (Model 13, AIC = 3350.7; sleep hours β = −0.02, p > 0.05; sleep quality β = −0.35, p < 0.001), and anticipated stress’s association was also fully attenuated (Model 14, AIC = 3361.8; stress β = 0.09, p > 0.05; enjoyment β = −0.24, p < 0.001) — a pattern distinct from the activity outcome, where anticipated stress retained marginal significance under the same comparison. A model combining all four contextual variables (Model 15, AIC = 3341.7) improved fit only modestly beyond either paired model. A reduced model retaining only sleep quality and anticipated enjoyment (Model 16, AIC = 3339.6) showed no significant loss of fit relative to Model 15 (*x*^2^ = 1.98, df = 2, p = 0.372), and in fact achieved a numerically lower AIC — further evidence that sleep hours and anticipated stress added no meaningful information beyond sleep quality and anticipated enjoyment.

Combining both predictor sets, a likelihood ratio test comparing Model 17 (all seven daily predictors, AIC = 2937.6) against Model 8 indicated that the contextual variables did not significantly improve model fit (*x*^2^ = 4.77, df = 4, p = 0.312). A further likelihood ratio test comparing Model 18 (the reduced contextual set combined with brain fog, forgetfulness, and mood, AIC = 2934.3) against Model 8 was also non-significant (*x*^2^ = 4.11, df = 2, p = 0.128); in Model 18, sleep quality (β = −0.10, p = 0.14) and anticipated enjoyment (β = −0.08, p = 0.23) did not reach significance. Model 17 vs. Model 18 showed no significant difference (*x*^2^ = 0.66, df = 2, p = 0.719). Because the difference in AIC between Model 8 (AIC = 2934.4) and Model 18 (AIC = 2934.3) was negligible (ΔAIC = 0.1), and neither sleep quality nor anticipated enjoyment reached significance in Model 18, Model 8 — the most parsimonious model, containing only brain fog, forgetfulness, and mood — was retained as the final model for this outcome.

In the final model (Model 8), brain fog (β = 1.16, SE = 0.07, OR = 3.19, p < 0.001), forgetfulness (β = 0.50, SE = 0.14, OR = 1.65, p < 0.001), and mood (β = 0.27, SE = 0.06, OR = 1.31, p < 0.001) were each independently and significantly associated with pain interference with concentration, while age, sex, and employment status showed no significant association.

### Comparison Across Outcomes

Across both outcomes, the model-building process followed a closely parallel trajectory. All seven daily predictors — brain fog, forgetfulness, mood, sleep hours, sleep quality, anticipated stress, and anticipated enjoyment — achieved statistical significance individually for both activity and concentration interference (Models 2–4 and 9–12), with brain fog consistently emerging as the single strongest individual predictor of either outcome (Model 2, AIC = 3097.7 for activity interference; AIC = 2962.7 for concentration interference). Within the primary predictor sequence, the fully combined model (Model 8) outperformed every individually or pairwise-specified alternative for both outcomes, and brain fog, forgetfulness, and mood each retained independent significance when modeled jointly. An identical pattern emerged within the contextual-variable sequence: sleep quality outperformed sleep hours, and anticipated enjoyment outperformed anticipated stress, both when compared individually (Models 9–12) and when evaluated jointly within their respective conceptually related pairs (Models 13–14). Sleep hours’ association was fully attenuated for both outcomes upon adjustment for sleep quality, while anticipated stress’s association was attenuated for both outcomes, remaining marginally significant for activity interference but fully attenuated for concentration interference. A reduced, two-variable contextual model (Model 16) demonstrated no significant loss of fit relative to the full four-variable contextual model (Model 15) for either outcome, substantiating the exclusion of sleep hours and anticipated stress from subsequent consideration in both cases.

The two outcomes diverged only at the final stage of model integration, when the contextual predictors (sleep quality, anticipated enjoyment) were combined with the primary predictors (brain fog, forgetfulness, mood). For pain interference with activity, this integration (Model 18) yielded a statistically significant improvement in fit relative to the primary-predictor-only model (Model 8; *x*^2^ = 15.26, df = 2, p < 0.001), with sleep quality and anticipated enjoyment each retaining independent significance in the combined model. For pain interference with concentration, the corresponding integration yielded no significant improvement in fit (*x*^2^ = 4.11, df = 2, p = 0.128), and neither sleep quality nor anticipated enjoyment attained significance once brain fog, forgetfulness, and mood were accounted for; the difference in AIC between Model 8 and Model 18 was negligible (ΔAIC = 0.1). Notably, brain fog’s association was also substantially stronger for concentration interference (OR = 3.19) than for activity interference (OR = 2.49) within each outcome’s respective final model. Accordingly, the final model selected differs between the two outcomes: Model 18, incorporating brain fog, forgetfulness, mood, sleep quality, and anticipated enjoyment, for pain interference with activity; and Model 8, incorporating brain fog, forgetfulness, and mood alone, for pain interference with concentration.

## Discussion

Migraine burden extends beyond headache itself to encompass cognitive and affective disturbance, including impaired concentration and emotional distress. Much of the existing evidence characterizing this burden derives from instruments such as the Migraine Disability Assessment Scale, which summarize functioning retrospectively over the preceding weeks or months and cannot capture how functional interference varies from one headache day to the next. Ecological momentary assessment offers a methodological corrective, capturing symptoms and their correlates close in time to the experience itself rather than reconstructing them from memory. The present study applied this approach in a smartphone-based migraine cohort to examine cognitive, affective, and contextual predictors, assessed concurrently with headache-related interference with daily activity and with concentration, on days when participants reported a headache since their last assessment, using cumulative link mixed models to accommodate the ordinal, unequally spaced structure of both outcomes.

Brain fog, forgetfulness, and mood disturbance constituted the dominant correlates of both outcomes, each remaining significant across every model of the primary predictor sequence, whereas age, sex, and employment status attained significance in none — indicating that functional interference on a headache day is governed more by concurrent symptom burden than by demographic or socioeconomic characteristics. Brain fog carried the largest effect size for both outcomes, with a stronger association for concentration than for activity interference, consistent with the closer conceptual link between subjective cognitive fog and difficulty concentrating specifically. Sleep quality and anticipated next-day enjoyment contributed meaningful explanatory value for activity interference beyond concurrent symptom burden alone, but not for concentration interference, indicating that these two facets of headache-related impairment, though correlated, are not governed by an identical set of daily influences.

These findings extend and, in some respects, qualify a growing body of ecological momentary assessment research on migraine-related functional and cognitive burden. Using a 28-day smartphone diary in women with migraine and comorbid obesity, Thomas and colleagues found that same-day pain intensity was a consistent predictor of pain interference across multiple domains, with older age and greater allodynia predicting elevated interference independent of pain intensity. The present findings depart from this pattern in an important respect: whereas Thomas and colleagues centered pain intensity as a primary predictor of interference, the present study was designed from the outset to examine cognitive, affective, and contextual correlates of interference on days when a headache was reported since the last assessment, and pain intensity was never considered as a candidate predictor. The two studies therefore address distinct, complementary questions rather than yielding directly conflicting results regarding the role of pain intensity.

Instead, brain fog, forgetfulness, and mood disturbance emerged as the dominant correlates of both outcomes, a pattern more consistent with Houtveen and Sorbi’s earlier ecological momentary assessment work identifying cognitive-affective functioning as a prominent feature of the migraine prodrome [1], and with cross-sectional evidence linking sleep disturbance and depressive symptoms to subjective cognitive complaints in migraine populations more broadly^23, 24^. Directly relevant to the present cohort, a companion analysis of the Migraine Impact on the Neurocognitive Dynamics (MIND)^11^ study modeled brain fog and forgetfulness as outcomes rather than predictors, using ordinal and logistic mixed-effects models, and found substantially higher odds of moderate-to-severe brain fog and same-day forgetfulness on headache days relative to headache-free days; it should be noted that this companion analysis defined headache days using the concurrent “right now” item rather than the “since the last assessment” item used in the present study, and drew on a slightly different analytic subsample (n = 139) within the same parent cohort. Considered alongside the present results, these two analyses of the same cohort suggest a complementary relationship: headache days are associated with markedly elevated cognitive and affective symptom burden, and that same symptom burden, in turn, is a strong correlate of how much a given headache day interferes with daily activity and concentration. Together, this body of evidence supports cognitive and affective symptoms, rather than pain intensity or demographic characteristics, as central and clinically actionable correlates of day-to-day functional impairment in migraine.

These findings carry direct implications for the clinical understanding and monitoring of headache-related functional impairment. Because brain fog, forgetfulness, and mood disturbance were more strongly and consistently associated with daily interference than any demographic or socioeconomic characteristic examined, clinical assessment focused narrowly on headache symptoms alone risks overlooking the factors most responsible for a patient’s day-to-day functional burden. Routine inquiry into same-day cognitive and affective symptoms, conducted alongside standard headache assessment, may therefore better capture the lived impact of migraine and identify patients for whom interventions targeting cognitive or affective symptoms specifically could meaningfully improve daily functioning.

The divergence between the two outcomes yields a further, more specific hypothesis: given that sleep quality and anticipated enjoyment were independently associated with activity interference but not with concentration, these contextual factors may constitute a distinct, modifiable target for improving activity-related functioning, though this observational association cannot establish a causal or interventional effect and would require prospective or experimental confirmation before informing treatment decisions.

These findings further underscore the value of the study’s underlying methodology, given that high-frequency, smartphone-based symptom monitoring captures within-person variability in migraine burden inaccessible to retrospective clinical assessment and thereby supports its continued application as a functional endpoint in future observational and treatment-evaluation research.

This study offers several methodological strengths. The use of ecological momentary assessment, rather than retrospective questionnaires, allowed same-day cognitive, affective, and contextual predictors to be examined in direct relation to functional interference on the same day, minimizing recall bias and enabling within-person analysis of a repeated-measures design. Modeling both outcomes as ordinal, rather than continuous, variables through cumulative link mixed models represent a further methodological strength, since this approach avoided the equal-interval assumption that a linear model would have imposed on a five-category response scale with unequal, data-dependent spacing between categories. The systematic, nested model-building strategy — evaluating each candidate predictor individually, in conceptually related pairs, and in combination, with model selection guided jointly by Akaike Information Criterion comparisons and likelihood ratio tests — provided a transparent and reproducible basis for identifying the final predictor set for each outcome, rather than relying on a single, unexamined model specification. This strategy further ensured that the most parsimonious model consistent with the data was retained for each outcome, as demonstrated by the selection of a three-predictor model for concentration interference once additional contextual variables failed to yield a meaningful improvement in fit. Finally, examining two related but distinct interference outcomes within the same cohort and analytic framework allowed genuine divergences between them, such as the differential contribution of sleep quality and anticipated enjoyment, to be identified with confidence rather than attributed to differences in sample or methodology across separate studies.

Several limitations warrant consideration. This study constitutes a secondary, exploratory, non-preregistered analysis of an observational cohort not originally designed for this specific research question, and its findings should accordingly be regarded as hypothesis-generating rather than confirmatory. Because all predictors and outcomes were derived from same-day self-report within a single daily survey, shared method variance may have contributed to the observed associations independent of any substantive underlying relationship. The eighteen-model comparison sequence conducted for each outcome was not accompanied by formal correction for multiple comparisons; although the nested, hypothesis-driven structure of these comparisons and the consistency of findings across both outcomes partially offset this concern, individual p-values warrant cautious interpretation. The within-day, observational design further precludes causal or temporal inference and cannot exclude reverse or bidirectional influence between symptom burden and functional interference. Several baseline subgroups, including Hispanic ethnicity and specific employment categories, comprised too few participants to support stable estimation, restricting inference across these characteristics; employment status was accordingly collapsed into a binary indicator, sacrificing clinically meaningful distinction for model stability. Finally, consistent with the MIND cohort, migraine diagnosis was self-reported rather than clinician-confirmed, and recruitment through digital platforms may have favored a more digitally engaged and socioeconomically advantaged sample, constraining the external validity of these findings.

Taken together, these findings position same-day cognitive and affective symptoms as the primary correlates of headache-related functional interference in this cohort, and support continued use of high-frequency digital monitoring to characterize and eventually intervene upon this burden in daily life.

## Conclusion

This study identified brain fog, forgetfulness, and mood disturbance as the dominant, consistent correlates of headache-related interference with activity and concentration in a smartphone-based migraine cohort, substantially exceeding the explanatory strength of demographic and socioeconomic characteristics. Sleep quality and anticipated next-day enjoyment provided additional predictive value for activity interference but not for concentration interference, indicating that these two domains of functional impairment are not fully explained by an identical set of daily influences. These findings support routine clinical assessment of cognitive and affective symptoms alongside conventional pain-focused evaluation and underscore the value of high-frequency digital monitoring for characterizing day-to-day migraine burden. Prospective, interventional research is needed to determine whether directly targeting these symptoms can reduce functional interference in daily life.

### Ethical Considerations

The study protocol was approved by the Shahid Beheshti University of Medical Science Institutional Review Board (IRB Protocol #000041), and all participants provided informed consent prior to enrollment. No additional ethical approval was required, as it involved only de-identified data provided under an institutional data-sharing agreement.

## Data Availability

The de-identified data used in this analysis are available from the corresponding author upon reasonable request and subject to institutional data-sharing agreements.

